# Characteristics and outcomes of patients with COVID-19 at high risk of disease progression receiving sotrovimab, oral antivirals or no treatment in Scotland

**DOI:** 10.1101/2023.06.09.23291195

**Authors:** Myriam Drysdale, Holly Tibble, Vishal Patel, Daniel C. Gibbons, Emily Lloyd, William Kerr, Calum Macdonald, Helen Birch, Aziz Sheikh

**Affiliations:** GSK, Brentford, Middlesex, UK; Usher Institute, University of Edinburgh, Scotland, UK

**Author notes:** **Corresponding author:** Myriam Drysdale, GSK House, 980 Great West Road, Brentford, London, TW8 9GS, UK. At the time of study (Vishal Patel).

**Keywords:** COVID-19, molnupiravir, monoclonal antibody, nirmatrelvir/ritonavir, Omicron BA.1, Omicron BA.2, Omicron BA.5, oral antivirals, sotrovimab

## Abstract

**Introduction:** Severe acute respiratory syndrome coronavirus 2 is constantly evolving. The clinical benefit of coronavirus disease 2019 (COVID-19) treatments against new circulating variants remains unclear. We sought to describe the real-world use of, and clinical outcomes associated with, early COVID-19 treatments among non-hospitalised patients with COVID-19 at highest risk of developing severe disease in Scotland.

**Methods:** Retrospective cohort study of non-hospitalised patients diagnosed with COVID-19 from 1 December 2021 to 25 October 2022, using administrative health data managed by Public Health Scotland and National Records of Scotland. Patients included in the study were aged ≥18 years, met at least one of the National Health Service highest-risk conditions criteria for early COVID-19 treatment, and had received outpatient treatment with sotrovimab, nirmatrelvir/ritonavir or molnupiravir, or no early COVID-19 treatment. Index date was defined as the earliest of either COVID-19-positive diagnosis or early COVID-19 treatment during the study period. Baseline patient characteristics and acute clinical outcomes in the 28 days following the index date were reported. To protect patient confidentiality, values of ≤5 were suppressed.

**Results:** A total of 2548 patients were included (492: sotrovimab, 276: nirmatrelvir/ritonavir, 71: molnupiravir, and 1709 eligible highest-risk untreated). Patients aged ≥75 years accounted for 6.9% (n=34/492) of the sotrovimab-treated group, 21.0% (n=58/276) of those treated with nirmatrelvir/ritonavir, 16.9% (n=12/71) of those treated with molnupiravir and 13.2% (n=225/1709) of untreated patients. Advanced renal disease was reported for 6.7% (n=33/492) of sotrovimab-treated and 4.7% (n=81/1709) of untreated patients, and five or fewer patients in the nirmatrelvir/ritonavir and molnupiravir cohorts. A high proportion of treated patients did not have a highest-risk condition reported in the database (71.7% for sotrovimab [n=353/492], 85.1% for nirmatrelvir/ritonavir [n=235/276], 85.9% for molnupiravir [n=61/71]). Five or fewer patients in each treated cohort experienced COVID-19-related hospitalisations during the 28-day acute period. For untreated patients, the percentage of COVID-19-related hospitalisations was 3.0% (n=48/1622). All-cause hospitalisations were experienced by 5.3% (n=25/476) of sotrovimab-treated patients, 6.9% (n=12/175) of nirmatrelvir/ritonavir-treated patients and 13.3% (n=216/1622) of untreated patients. Five or fewer patients in the molnupiravir cohort experienced all-cause hospitalisation. There were no deaths within 28 days of index for patients in the treated cohorts. Mortality was 4.3% (n=70/1622) in untreated patients (18.6% [n=13/70] had COVID-19 as the primary cause). In our analyses of outcomes for sotrovimab-treated and untreated patients during BA.1, BA.2 and BA.5 predominance, COVID-19-related hospitalisation rates were consistent, with n≤5 for sotrovimab-treated patients in each period.

**Conclusions:** Our findings indicate that sotrovimab was often used amongst patients who were aged <75 years old and had advanced renal disease. Among patients who received early COVID-19 treatment, proportions of all-cause hospitalisation and death within 28 days of treatment were low.

## Introduction

Coronavirus disease 2019 (COVID-19) is caused by infection with severe acute respiratory syndrome coronavirus 2 (SARS-CoV-2). The rapid global spread of SARS-CoV-2 resulted in the declaration of a pandemic by the World Health Organization in March 2020.^1^ Older individuals, immunocompromised patients or those with comorbidities such as cancer, diabetes, advanced renal disease, chronic obstructive pulmonary disease or cardiovascular disease are known to be at increased risk of developing severe COVID-19, which may result in hospitalisation or death.^2,3^

In the United Kingdom (UK), early COVID-19 treatment with either antivirals (nirmatrelvir/ritonavir and molnupiravir) or monoclonal antibodies (mAbs; sotrovimab) is recommended for people with ‘highest-risk’ conditions. At the time of this study, these included solid cancer, advanced renal disease, advanced liver disease and human immunodeficiency virus (HIV)/ acquired immune deficiency syndrome (AIDS).^4^

Sotrovimab is a dual-action engineered human IgG1κ mAb derived from the parental mAb S309, a potent neutralising mAb directed against the spike protein of SARS-CoV-2.^5-8^ Sotrovimab administered as a single 500-mg intravenous dose was shown in COMET-ICE (NCT04545060), a randomised clinical trial (conducted from August 2020 to March 2021, when the original ‘wild-type’ variant was predominant), to significantly reduce the relative risk of all-cause >24-hour hospitalisation or death due to any cause by 79% compared with placebo in high-risk patients with mild-to-moderate COVID-19.^9^ In December 2021, sotrovimab received conditional marketing authorisation in the UK for use in symptomatic adults and adolescents (aged ≥12 years and weighing ≥40 kg) with acute COVID-19 infection who did not require supplemental oxygen and were deemed to be at increased risk of progression to severe COVID-19.^10^

Two oral antivirals, molnupiravir and nirmatrelvir/ritonavir, have been shown to reduce the risk of progression to severe COVID-19 compared with placebo among high-risk patients with mild-to-moderate COVID-19.^11-13^ Molnupiravir and nirmatrelvir/ritonavir received conditional marketing authorisation in November 2021 and December 2021, respectively, for use in patients with COVID-19 who were deemed to be at increased risk of progression to severe COVID-19.^14,15^

During the study period (December 2021 to October 2022), National Health Service (NHS) clinical guidelines recommended sotrovimab and nirmatrelvir/ritonavir as first-line treatment options, and molnupiravir as a third-line option.^16^ It should be noted that nirmatrelvir/ritonavir is contraindicated for many highest-risk patients, including those with advanced renal disease, HIV/AIDS and certain types of cancer.^17^ These guidelines apply to all parts of the UK, including Scotland.^18^

Here, we describe real-world use of early COVID-19 treatments (including patient characteristics and clinical outcomes) for the management of non-hospitalised patients with COVID-19 at highest risk of developing severe disease in Scotland. The descriptive nature of this study without formal statistical comparison was intended to inform the feasibility of a potential comparative effectiveness analysis of sotrovimab versus no early COVID-19 treatment.

## Methods

### Study design and data source

This retrospective cohort study followed STROBE and RECORD reporting guidelines. We used data from administrative health datasets managed by Public Health Scotland and National Records of Scotland, linked and pseudonymised by the electronic Data Research and Innovation Service.

The study cohort was drawn from the Scottish general practitioner-registered population living within six health boards (i.e. Ayrshire & Arran, Dumfries & Galloway, Forth Valley, Greater Glasgow & Clyde, Lanarkshire and Lothian) that utilised the Hospital Electronic Prescribing and Medicines Administration (HEPMA) system for recording the administration and prescription of COVID-19 therapies.

The index date (Day 1) was defined as the earliest date of a confirmed COVID-19 diagnosis (via reverse transcriptase polymerase chain reaction or lateral flow test [LFT]) or treatment date during the study period. Patients were followed-up for 28 days (Day 28) from the index date, which was defined as the acute period, during which patient outcomes were evaluated. The study included patients diagnosed with COVID-19 from 1 December 2021 to 25 October 2022. The baseline period was defined as the two years prior to index for secondary care events, and one year prior to index for general practitioner prescriptions.

This study received data access and processing approval from the Public Benefit and Privacy Panel for Health and Social Care, NHS Scotland on 9 November 2022. We complied with all applicable laws regarding subject privacy. No direct subject contact or primary collection of individual human subject data occurred as part of this study. Study results were reported in tabular form and aggregate analyses that omitted subject identification.

Publications, reports or any other research outputs do not include subject identifiers and low patient numbers (n≤5) were suppressed.

### Study population

Non-hospitalised patients were eligible for inclusion if they were aged ≥18 years on the index date; had a COVID-19 diagnosis/positive SARS-CoV-2 polymerase chain reaction or LFT; lived within one of the six geographical zones attached to a Scottish health board which used the HEPMA prescribing system; met at least one of the NHS highest-risk conditions criteria for receiving early treatment with sotrovimab, nirmatrelvir/ritonavir or molnupiravir (as defined by the presence of diagnosis codes) (Table 1); and received outpatient treatment with sotrovimab, nirmatrelvir/ritonavir or molnupiravir, or received no early COVID-19 treatment.

**Table 1.**
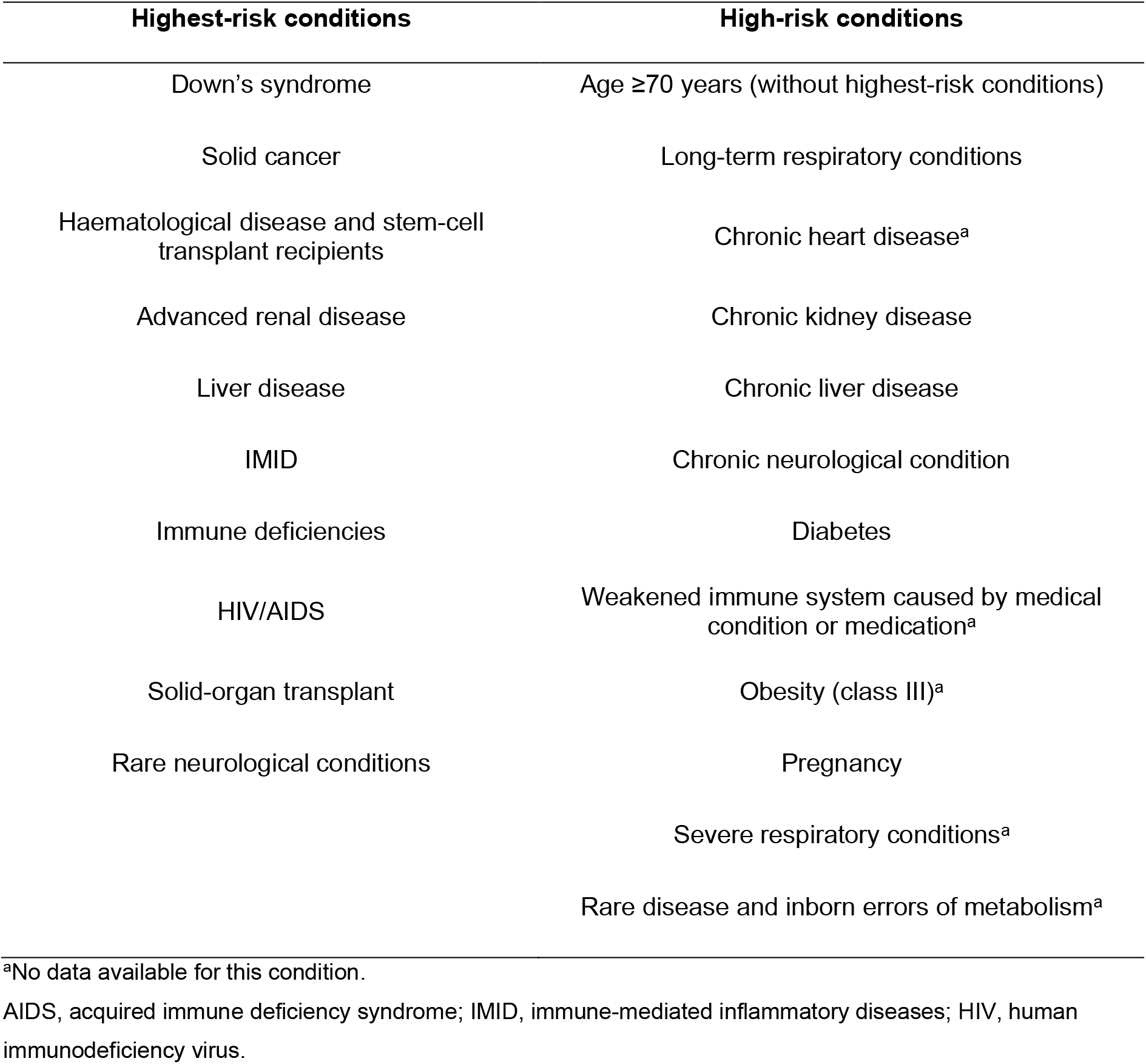
Highest- and high-risk conditions criteria

| Highest-risk conditions | High-risk conditions |
| --- | --- |
| Down's syndrome | Age ≥70 years (without highest-risk conditions) |
| Solid cancer | Long-term respiratory conditions |
| Haematological disease and stem-cell transplant recipients | Chronic heart disease<sup>a</sup> |
| Advanced renal disease | Chronic kidney disease |
| Liver disease | Chronic liver disease |
| IMID | Chronic neurological condition |
| Immune deficiencies | Diabetes |
| HIV/AIDS | Weakened immune system caused by medical condition or medication<sup>a</sup> |
| Solid-organ transplant | Obesity (class III)<sup>a</sup> |
| Rare neurological conditions | Pregnancy |
|  | Severe respiratory conditions<sup>a</sup> |
|  | Rare disease and inborn errors of metabolism<sup>a</sup> |

<sup>a</sup>No data available for this condition.
AIDS, acquired immune deficiency syndrome; IMID, immune-mediated inflammatory diseases; HIV, human immunodeficiency virus.

At the time of the study, the NHS highest-risk criteria included Down’s syndrome, solid cancer, haematological diseases (including cancers), advanced renal disease, advanced liver disease, immune-mediated inflammatory disorders (IMID), immune deficiencies, HIV/AIDS, solid-organ and stem-cell transplant recipients and rare neurological conditions.^4^ These highest-risk criteria were evaluated during the baseline period.

Patients were excluded if they received more than one COVID-19 treatment (sotrovimab, nirmatrelvir/ritonavir, molnupiravir or remdesivir) in an outpatient setting during the acute period; they received remdesivir as an early treatment in an outpatient setting; or they received sotrovimab, nirmatrelvir/ritonavir or molnupiravir while in an inpatient setting (defined as overnight admission on the day of or prior to treatment, and discharge after the day of treatment).

### Study outcomes

The primary outcomes of this study were the proportions of patients with all-cause and COVID-19-related hospitalisations during the acute period (28 days following index). COVID-19-related hospitalisations were defined as any non-elective hospital visit for which COVID-19 was listed in the primary diagnosis field (amongst patients in which the hospitalisation episode was complete, i.e. discharge had occurred and clinical coding was complete).

Secondary outcomes included the number of all-cause and COVID-19-related inpatient hospitalisation days, the proportion of patients with a critical care admission as part of hospitalisation, the proportion of patients requiring non-invasive ventilation and mechanical ventilation, and the proportion of deaths during the acute period.

For all outcomes, patients were excluded if they had less than 45 days between the index diagnosis and data extraction censoring dates (regardless of whether they died in this interval) for the outcomes data (6 October 2022).

Patient characteristics were also recorded, including age, sex, COVID-19 vaccination status and comorbidity history. COVID-19 vaccination data were only available from 1 December 2021; any vaccinations before this date were therefore unknown, and individuals were classified as having ‘missing’ time since last vaccination. Cohorts were described in relation to ‘highest-risk’ conditions that made patients eligible for early treatment with sotrovimab, nirmatrelvir/ritonavir or molnupiravir, as mentioned above. Additionally, the cohorts were described in relation to other ‘high-risk’ conditions that may predispose patients to severe COVID-19 outcomes (Table 1).

Outcomes were reported for the following cohorts: Cohort 1, patients receiving early treatment with sotrovimab; Cohort 2, patients receiving early treatment with nirmatrelvir/ritonavir; Cohort 3, patients receiving early treatment with molnupiravir; and Cohort 4, patients at highest risk who received no early COVID-19 treatment.

A subgroup analysis was also conducted. We described 28-day COVID-19-related hospitalisation among sotrovimab-treated patients (Cohort 1) and those without any early COVID-19 treatments (Cohort 4) during the periods of Omicron BA.1 (1 December 2021–28 February 2022), Omicron BA.2 (1 March–31 May 2022) and Omicron BA.5 (1 June–30 September 2022) subvariant predominance in the UK (Figure 1).^19^ These analyses were not performed for patients treated with nirmatrelvir/ritonavir or molnupiravir due to the low sample sizes expected in these cohorts. Due to low rates of sequencing, the time periods of most prevalent circulating variants were used as a proxy for the infecting viral variant in patients diagnosed with COVID-19.

**Figure 1.**
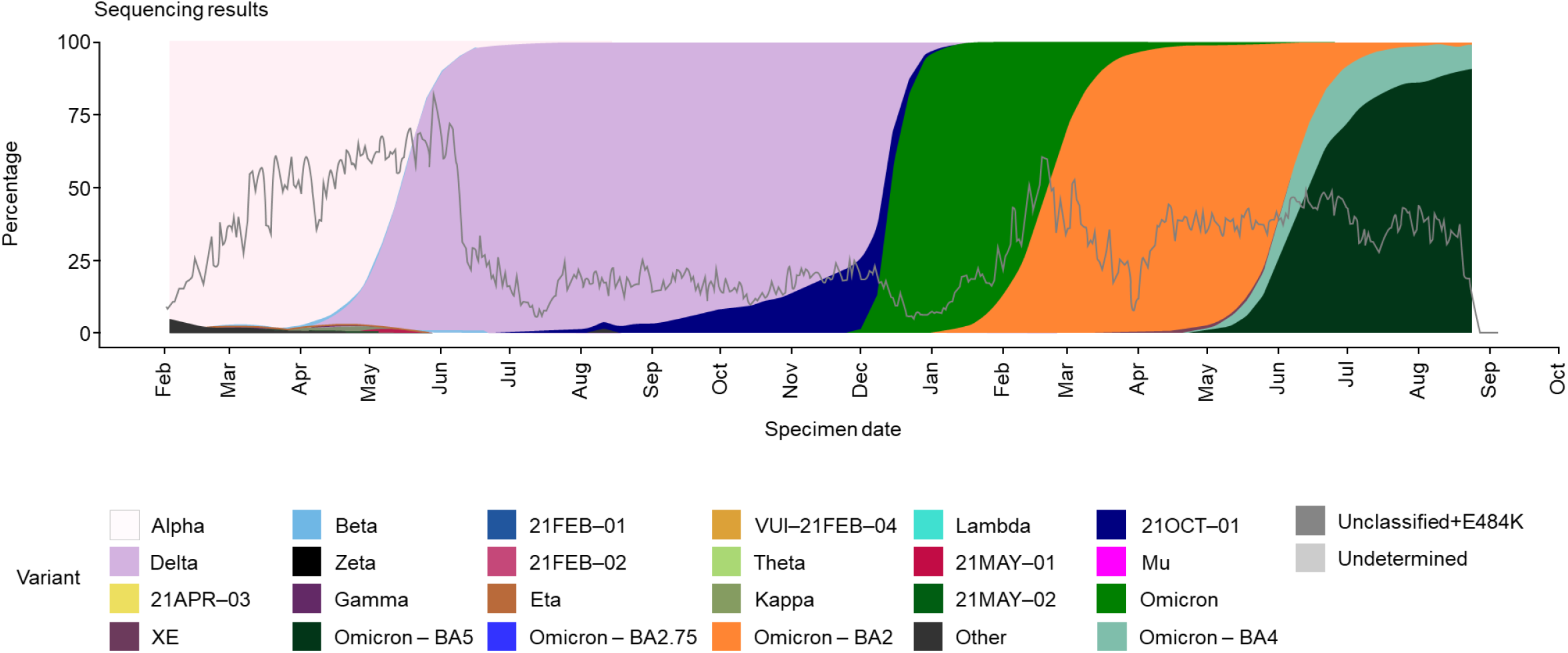
SARS-CoV-2 variant predominance from February 2021 to September 2022 UK Health Security Agency. SARS-CoV-2 variants of concern and variants under investigation in England: Technical briefing 45 (2022). Reprinted from: https://assets.publishing.service.gov.uk/government/uploads/system/uploads/attachment_data/file/1115071/Technical-Briefing-45-9September2022.pdf. Contains public sector information licensed under the Open Government Licence v3.0: https://www.nationalarchives.gov.uk/doc/open-government-licence/version/3/. Accessed 23 May 2023. SARS-CoV-2, severe acute respiratory syndrome coronavirus 2.

### Data analysis

Continuous variables (e.g. age) were summarised using mean, standard deviation, median, interquartile range and range. Categorical variables (e.g. sex) were described using frequencies and percentages. Due to information governance and data suppression rules used in the study, counts of between 0 and 5 were suppressed and are reported as n≤5 throughout (unless they were structural zeros, caused by inclusion/exclusion criteria). This suppression did not apply to mortality data.

## Results

### Patient demographics and baseline characteristics

Following application of the inclusion and exclusion criteria, demographics and baseline characteristics were available for 2548 patients, including 492 patients treated with sotrovimab, 276 patients treated with nirmatrelvir/ritonavir, 71 patients treated with molnupiravir and 1709 eligible highest-risk untreated patients (Figure 2). Baseline characteristics are reported in Table 2.

**Table 2.** Patient characteristics

|  | **Sotrovimab**<br>**(n=492)** | **Nirmatrelvir/**<br>**ritonavir**<br>**(n=276)** | **Molnupiravir**<br>**(n=71)** | **Untreated**<br>**(n=1709)** |
| --- | --- | --- | --- | --- |
| **Age (years)** |  |  |  |  |
| Mean (SD) | 54.6 (15.0) | 60.9 (17.3) | 59.6 (14.0) | 55.1 (17.9) |
| Median (IQR) | 55 (44–65) | 62 (50–73) | 60 (49.5–69.5) | 57 (41–69) |
| **Age group (years), n (%)** |  |  |  |  |
| 18–54 | 242 (49.2) | 95 (34.4) | 22 (31.0) | 783 (45.8) |
| 55–64 | 121 (24.6) | 61 (22.1) | 20 (28.2) | 369 (21.6) |
| 65–74 | 95 (19.3) | 62 (22.5) | 17 (23.9) | 332 (19.4) |
| ≥75 | 34 (6.9) | 58 (21.0) | 12 (16.9) | 225 (13.2) |
| **Female sex, n (%)** | 286 (58.1) | 164 (59.4) | 42 (59.2) | 949 (55.5) |
| **Number of vaccinations,**<br>**n (%)** |  |  |  |  |
| 0–2 | 21 (4.3) | 19 (6.9) | ≤5 | 315 (18.4) |
| 3 | 281 (57.1) | 126 (45.7) | 41 (57.7) | 1036 (60.6) |
| 4 | 160 (32.5) | 88 (31.9) | 23 (32.4) | 283 (16.6) |
| ≥5 | 30 (6.1) | 43 (15.6) | ≤5 | 75 (4.4) |
| **Time since last**<br>**vaccination (days), n (%)** |  |  |  |  |
| 1–60 | 164 (33.3) | 37 (13.4) | 17 (23.9) | 369 (21.6) |
| 61–120 | 204 (41.5) | 56 (20.3) | 28 (39.4) | 542 (31.7) |
| 121–180 | 74 (15.0) | 88 (31.9) | 10 (14.1) | 371 (21.7) |
| ≥181 | 50 (10.2) | 95 (34.4) | 16 (22.5) | 427 (25.0) |
| **Highest-risk conditions, n (%)<sup>a</sup>** |  |  |  |  |
| Solid cancer | 45 (9.1) | 16 (5.8) | ≤5 | 501 (29.3) |
| Haematological<br>disease, sickle cell<br>disease and stem-cell<br>transplant recipients | 28 (5.7) | 15 (5.4) | ≤5 | 89 (5.2) |
| Advanced renal disease | 33 (6.7) | ≤5 | ≤5 | 81 (4.7) |
| Liver disease | ≤5 | ≤5 | ≤5 | 23 (1.3) |
| IMiD | 31 (6.3) | 12 (4.3) | ≤5 | 964 (56.4) |
| Immune deficiencies | ≤5 | ≤5 | ≤5 | 7 (0.4) |
| HIV/AIDS | ≤5 | ≤5 | ≤5 | ≤5 |
| Solid-organ transplant | ≤5 | ≤5 | ≤5 | ≤5 |
| Rare neurological<br>conditions | 9 (1.8) | ≤5 | ≤5 | 72 (4.2) |
| **Patients with no highest-**<br>**risk diagnoses for**<br>**developing COVID-19**<br>**complications, n (%)** | 353 (71.7) | 235 (85.1) | 61 (85.9) | 0 |
| **High-risk conditions, n (%)<sup>b</sup>** |  |  |  |  |
| Age ≥70 years (and no<br>highest-risk conditions) | 56 (11.4) | 84 (30.4) | 17 (23.9) | 0 |
| Chronic respiratory<br>conditions | 15 (3.0) | 8 (2.9) | ≤5 | 49 (2.9) |
| Other chronic kidney disease | ≤5 | ≤5 | ≤5 | ≤5 |
| Other chronic liver disease | ≤5 | ≤5 | ≤5 | 8 (0.5) |

Due to information governance and data suppression rules used in the study, counts of between 0 and 5 were suppressed and are reported as n≤5 throughout, excluding structural zeros (due to inclusion/exclusion criteria).
<sup>a</sup>No cases of Down's syndrome were identified from the available data; <sup>b</sup>No cases of pregnancy, chronic moderate-risk neurological conditions or diabetes were identified from the available data.
AIDS, acquired immune deficiency syndrome; COVID-19, coronavirus disease 2019; HIV, human immunodeficiency virus; IMID, immune-mediated inflammatory disease; IQR, interquartile range; SD standard deviation.

**Figure 2.**
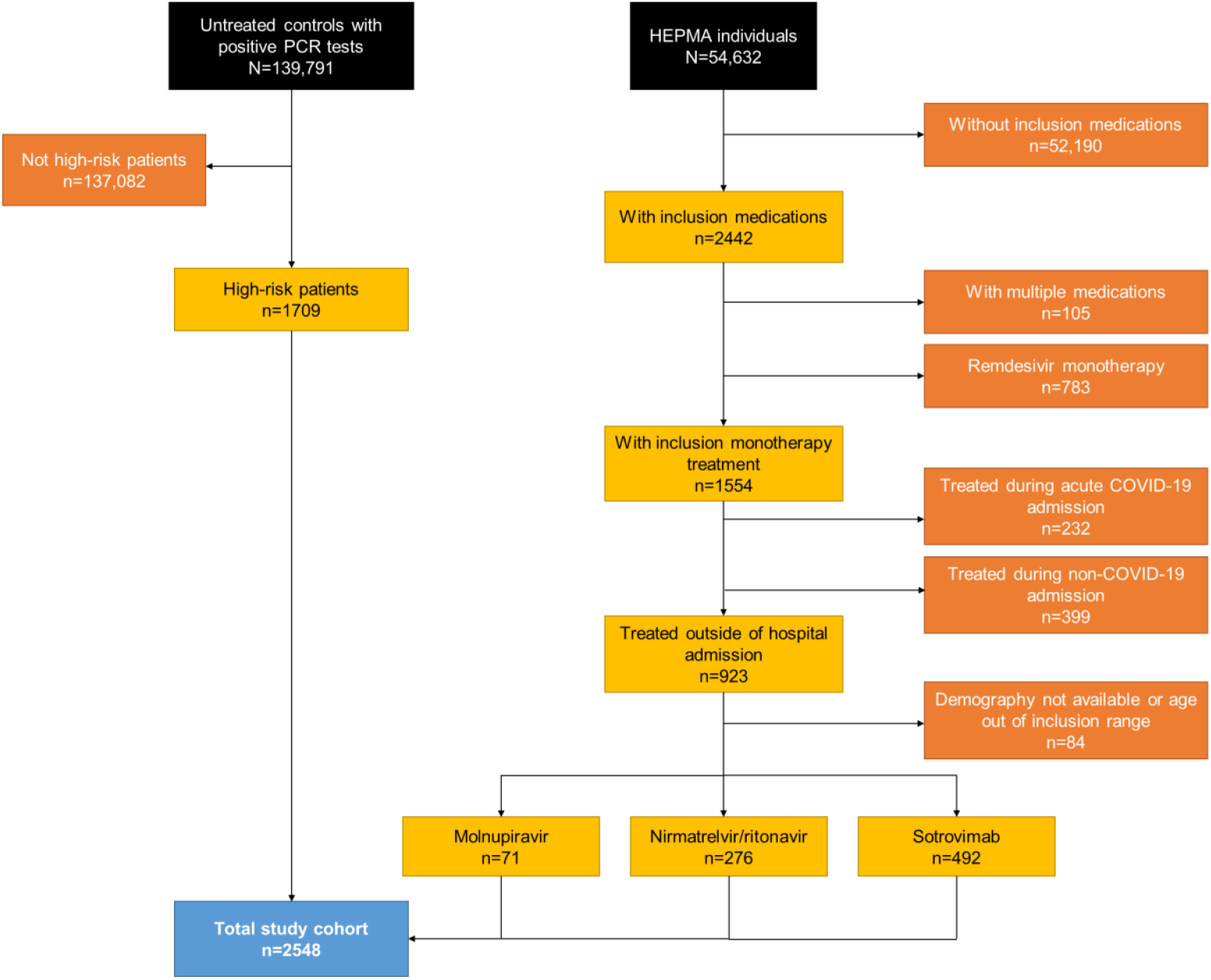
Flow diagram of patient cohort inclusion/exclusion criteria COVID-19, coronavirus disease 2019; HEPMA, Hospital Electronic Prescribing and Medicines Administration; PCR, polymerase chain reaction.

Patients aged ≥75 years accounted for 6.9% (n=34/492) of the sotrovimab-treated group, 21.0% (n=58/276) of those treated with nirmatrelvir/ritonavir, 16.9% (n=12/71) of those treated with molnupiravir and 13.2% (n=225/1709) of untreated patients.

A high proportion of patients treated with an early COVID-19 treatment did not have an identifiable highest-risk condition reported in the database (71.7% for sotrovimab [n=353/492], 85.1% for nirmatrelvir/ritonavir [n=235/276], 85.9% for molnupiravir [n=61/71] and 0% for untreated patients [resulting from the inclusion criteria in this group]). A high percentage of untreated patients had solid cancer (29.3%, n=501/1709) and IMID (56.4%, n=964/1709), while comparatively lower percentages were reported for other highest-risk comorbidities among this cohort (Table 2). Of patients treated with sotrovimab, 9.1% (n=45/492) had solid cancer and 6.3% (n=31/492) had IMID. Solid cancer was the most frequently reported highest-risk condition for patients treated with nirmatrelvir/ritonavir (5.8%, n=16/276), and each of the highest-risk comorbidities were reported for five or fewer patients treated with molnupiravir. Advanced renal disease was reported for 6.7% (n=33/492) of patients treated with sotrovimab and 4.7% (n=81/1709) of untreated patients. Five or fewer patients treated with nirmatrelvir/ritonavir (n=≤5/276) and molnupiravir (n=≤5/71) had advanced renal disease.

### Acute-period outcomes

In total, 8.5% (n=216/2548) of patients did not have sufficient observation time (45 days, including after death, from index diagnosis to data extraction date) to assess their acute-period outcomes (sotrovimab: n=16/492 [3.3%]; nirmatrelvir/ritonavir: n=101/276 [36.6%]; molnupiravir: n=12/71 [16.9%]; untreated: n=87/1709 [5.1%]).

The percentage of patients who experienced an all-cause hospitalisation was 5.3% (n=25/476) for sotrovimab-treated patients and 6.9% (n=12/175) for nirmatrelvir/ritonavir-treated patients (Table 3). Five or fewer patients treated with molnupiravir (n=≤5/59) experienced an all-cause hospitalisation. For untreated patients, the percentage of all-cause hospitalisations was 13.3% (n=216/1622). The median (interquartile range) number of days in hospital due to any cause during the acute period was 5.0 (2–9) for sotrovimab-treated patients, 3.5 (2–9.5) for nirmatrelvir/ritonavir and 6.0 (3–14) for untreated patients. The median length of stay could not be reported for molnupiravir due to low patient numbers.

**Table 3.** Acute period all-cause and COVID-19-related outcomes

|  | **Sotrovimab**<br>**(n=476)** | **Nirmatrelvir/**<br>**ritonavir**<br>**(n=175)** | **Molnupiravir**<br>**(n=59)** | **Untreated**<br>**(n=1622)** |
| --- | --- | --- | --- | --- |
| **All-cause** |  |  |  |  |
| Inpatient (overnight) hospitalisation, n (%) | 25 (5.3) | 12 (6.9) | ≤5 | 216 (13.3) |
| Inpatient hospitalisation days |  |  |  |  |
| Mean (SD) | 7.0 (6.0) | 8.8 (11.1) | NR | 10.8 (14.0) |
| Median (IQR) | 5 (2–9) | 3.5 (2–9.5) |  | 6 (3–14) |
| Critical care admission, n (%) | ≤5 | ≤5 | ≤5 | 10 (0.6) |
| Non-invasive ventilation, n (%) | ≤5 | ≤5 | ≤5 | ≤5 |
| Mechanical ventilation, n (%) | ≤5 | ≤5 | ≤5 | ≤5 |
| Inpatient (overnight) hospitalisation with COVID-19 recorded in any diagnosis field, n (%) | ≤5 | ≤5 | ≤5 | 66 (4.1) |
| Deaths, n (%) | 0 | 0 | 0 | 70 (4.3) |
| **COVID-19-related** |  |  |  |  |
| Inpatient (overnight) hospitalisation (primary diagnosis), n (%) | ≤5 | ≤5 | ≤5 | 48 (3.0) |
| Inpatient hospitalisation days |  |  |  |  |
| Mean (SD) | NR | NR | NR | 8.4 (7.4) |
| Median (IQR) |  |  |  | 6 (3–12) |
| Critical care admission, n (%) | ≤5 | ≤5 | ≤5 | ≤5 |
| Non-invasive ventilation, n (%) | ≤5 | ≤5 | ≤5 | ≤5 |
| Mechanical ventilation,<br>n (%) | ≤5 | ≤5 | ≤5 | ≤5 |
| Deaths, n (%) | 0 | 0 | 0 | 13 (0.8) |

The hospitalisations dataset ran from 1 December 2019 to 11 October 2022, the intensive care unit dataset ran from 1 December 2021 to 23 October 2022 and the deaths dataset ran from 2 December 2021 to 6 October 2022. These three outcomes' datasets were censored to align with the earliest dataset (to 6 October 2022) for consistency.
Due to information governance and data suppression rules used in the study, counts of between 0 and 5 were suppressed and are reported as n≤5 throughout, except for mortality data, which does not require suppression. COVID-19, coronavirus disease 2019; IQR, interquartile range; NR, not reported; SD, standard deviation.

Five or fewer patients in all cohorts required non-invasive ventilation or mechanical ventilation. Critical care admissions were experienced by five or fewer patients in each of the treated cohorts, and by 10 (0.6%) untreated patients.

Five or fewer patients treated with each of sotrovimab, nirmatrelvir/ritonavir and molnupiravir experienced COVID-19-related hospitalisations during the 28-day acute period (Table 3). For untreated patients, the percentage of COVID-19-related hospitalisations was 3.0% (n=48/1622). Five or fewer patients in all cohorts required non-invasive ventilation or mechanical ventilation, or experienced critical care admission.

There were no deaths within 28 days of the index date for patients treated with sotrovimab, nirmatrelvir/ritonavir or molnupiravir. Mortality was 4.3% (n=70/1622) in untreated patients, of whom 13 (18.6%) had COVID-19 as the primary cause (Table 3).

### COVID-19-related hospitalisations during periods of BA.1, BA.2 and BA.5 predominance

During the BA.1 predominance period, 283 patients were treated with sotrovimab and 721 patients received no treatment (Table 4). Similar to the overall analysis, five or fewer patients treated with sotrovimab (n=≤5/283) experienced a COVID-19-related hospitalisation. The percentage for untreated patients was 3.1% (n=22/721).

**Table 4.** Acute period COVID-19-related hospitalisations during periods of Omicron BA.1, BA.2 and BA.5 predominance

|  | **Sotrovimab** | **Untreated** |
| --- | --- | --- |
| **BA.1** (1 December 2021 to 28 February 2022) | ≤5/283 | 22/721 (3.1%) |
| **BA.2** (1 March 2022 to 31 May 2022) | ≤5/144 | 14/625 (2.2%) |
| **BA.5** (1 June 2022 to 30 September 2022) | ≤5/49 | 12/276 (4.3%) |

Due to information governance and data suppression rules used in the study, counts of between 0 and 5 were suppressed and are reported as n≤5 throughout.
COVID-19, coronavirus disease 2019.

During the BA.2 predominance period, 144 patients were treated with sotrovimab and 625 did not receive any treatment (Table 4). Five or fewer patients treated with sotrovimab (n=≤5/144) and 2.2% (n=14/625) of untreated patients had COVID-19-related hospitalisations.

During BA.5 variant predominance, 49 patients were treated with sotrovimab and 276 patients received no treatment (Table 4). Five or fewer sotrovimab-treated patients (n=≤5/49) and 4.3% (n=12/276) of untreated patients had COVID-19-related hospitalisations.

## Discussion

This study described the characteristics and severe clinical outcomes of non-hospitalised patients who received early treatment for COVID-19 in Scotland from 1 December 2021 to 25 October 2022, or those who may have been eligible but did not receive treatment. Our findings indicate that sotrovimab was used amongst patients who are slightly younger (6.9% were aged ≥75 years versus 21.0% for nirmatrelvir/ritonavir and 16.9% for molnupiravir), mainly with advanced renal disease. We also found that patients who received early COVID-19 treatment with sotrovimab or antivirals experienced low levels of hospitalisation during the 28 days following treatment administration. Low levels of death due to any cause were also observed. The proportions of patients with severe clinical outcomes were, however, higher for patients who did not receive an early COVID-19 treatment. Across subvariant predominance periods (BA.1, BA.2 and BA.5), five or fewer sotrovimab-treated patients experienced COVID-19-related hospitalisations.

The observed characteristics of patients treated with sotrovimab differed to those described in a recent real-world study on early treatments for COVID-19 conducted in another part of the UK.^20^ In a retrospective cohort study of non-hospitalised patients who received early treatment for, or were diagnosed with, COVID-19 in North West London between 1 December 2021 and 31 May 2022, Patel *et al*. reported descriptive results for patients treated with sotrovimab, nirmatrelvir/ritonavir or molnupiravir. Sotrovimab was found to be utilised most often amongst older patients with multiple comorbidities that increased their risk of severe COVID-19, such as advanced renal disease.^20^ In the present study, we found that patients treated with sotrovimab were younger than the other treated cohorts, but also more likely to have advanced renal disease. Of note, we reported that a particularly high number of untreated patients had IMID (56.4%). This indicates that the effective eligibility criteria were likely narrower than the definitions used herein, with certain IMIDs associated with higher risk and real-world treatment allocation being reflective of this.

In the current study, the proportion of all-cause hospitalisations was low for patients treated with sotrovimab and antivirals. These results are supported by those of other recent real-world studies. Patel *et al*. reported low all-cause hospitalisations among patients treated with sotrovimab, nirmatrelvir/ritonavir and molnupiravir, with low hospitalisation rates for sotrovimab being consistent amongst patients with advanced renal disease, those aged 18– 64 and ≥65 years, and across periods of Omicron BA.1, BA.2 and BA.5 predominance (through July 2022).^20^ Our results for the overall cohort are also similar to those of a recent retrospective cohort study of patients presumed to be treated with sotrovimab in England, which reported that 4.6% of patients experienced an all-cause hospitalisation.^21^

A recent publication on the effectiveness of sotrovimab versus molnupiravir in Scotland and England investigated outcomes among patients on kidney replacement therapy using the OpenSAFELY platform and linked data from the UK Renal Registry. Among this patient population, sotrovimab treatment was associated with a lower risk of severe COVID-19 outcomes compared with molnupiravir (hazard ratio 0.35, 95% confidence interval [CI] 0.17, 0.71).^22^ In another publication on the comparative effectiveness of sotrovimab versus molnupiravir in England, again conducted using the OpenSAFELY platform, sotrovimab treatment was associated with a significant reduction in risk of severe COVID-19 outcomes in comparison to treatment with molnupiravir during periods of BA.1 (primary analysis) and BA.2 (exploratory analysis) predominance.^23^ In an additional study, sotrovimab was found to offer a similar level of protection against disease progression in patients with COVID-19 infected with Omicron BA.1 and BA.2.^24^ Similarly, in an observational study in the United States, treatment with sotrovimab was associated with a reduced risk of all-cause hospitalisation and mortality during periods when early Omicron variants were predominant.^25^ In a further study conducted using the OpenSAFELY platform, no substantial differences in the risk of severe COVID-19 outcomes were observed between patients treated with sotrovimab and nirmatrelvir/ritonavir from February–October 2022, when BA.2 and BA.5 were predominant.^26^ In another recent study, the OpenSAFELY platform was used to emulate target trials to estimate the effectiveness of sotrovimab or molnupiravir versus no treatment during periods of BA.1 and BA.2 predominance. Estimated hazard ratios for 28-day COVID-19-related hospitalisation or death were 0.76 (95% CI 0.66, 0.89) during BA.1 and 0.92 (95% CI 0.79, 1.06) during BA.2 for sotrovimab versus no treatment.^27^

We previously assessed the uptake of mAbs and antiviral therapies in Scotland, and discussed whether these treatments were being used as recommended.^28^ We reported that only around half of eligible patients received a mAb/antiviral for COVID-19, but that the vast majority of patients who received treatment did so within the recommended timeframe. In this study, we also found that some eligible patients (based on COVID-19 diagnosis and high-risk comorbidities) were not treated; the reasons for this were not ascertained as part of this study but is of interest for future research.

As discussed previously, five or fewer sotrovimab-treated patients experienced COVID-19-related hospitalisations across periods of BA.1, BA.2 and BA.5 predominance. Due to data suppression rules and small patient numbers (particularly with decreasing sotrovimab use during BA.5), interpretation of these results is challenging, particularly during periods of BA.2 and BA.5 predominance. The risk of ecological bias should be noted, as we did not have access to sequencing data to confirm the variant for each patient.

It should also be noted that these descriptive results require confirmation with formal statistical testing adjusting for confounding factors. Due to suppressed numbers resulting from low event rates and small sample sizes, and the high number of patients with no diagnoses associated with highest-risk of developing COVID-19 complications, future formal comparison was not considered feasible.

This study has several limitations which should be considered. Firstly, the study presents descriptive analyses with no adjustment for differences in patient characteristics between cohorts, meaning that results may be subject to bias and confounding. Further, the limited sample size (particularly in the molnupiravir cohort) led to frequent suppression of small values, which limits the comparability and interpretability of the results, and may introduce bias. The high proportion of patients with missing highest-risk comorbidities in the treated groups further reduces the feasibility of formal comparison adjusting for confounders. We did not collect data on COVID-19 severity and symptoms seen at onset of disease, and therefore cannot confirm that the cohorts were comparable in this regard. It is more likely that the sotrovimab-treated cohort had mild-to-moderate COVID-19, while some of those who were untreated were asymptomatic, mildly symptomatic or improving. In addition, 85% of the treated cohort had no identified reverse transcriptase polymerase chain reaction or (registered) LFT in the 40 days prior to treatment, whereas the untreated cohort were required to have a positive test recorded. These ‘untested’ treated patients were likely to have tested at home and not reported their results. There were limited sequencing data available for patients included in the study, and dominance period for Omicron subvariant was instead used as a surrogate. The proportion of patients without highest-risk conditions was also high across the treated cohorts due to missing data. Finally, comorbidities were estimated using only inpatient admissions and procedures data. As such, conditions treated primarily in specialist departments (including maternity wards and renal wards) or in primary care were not possible to ascertain.

## Conclusions

Our findings indicate that among patients who received early COVID-19 treatment with sotrovimab or antivirals in Scotland, low proportions experienced all-cause hospitalisations and death within 28 days of treatment. Sotrovimab was observed to be frequently utilised in patients with advanced renal disease and those aged below 75 years old in Scotland.

## Data Availability

Data for this study (ID 2223-0033) are held by the National Services Scotland electronic Data Research and Innovation Service in the National Safe Haven. Restrictions apply to the availability of these data, which were used under license for the current study, and so are not publicly available. Data would be made available from a reasonable request to.

## Medical writing, editorial and other assistance

Editorial support (in the form of writing assistance, including preparation of the draft manuscript under the direction and guidance of the authors, collating and incorporating authors’ comments for each draft, assembling tables, grammatical editing and referencing) was provided by Kathryn Wardle of Apollo, OPEN Health Communications, in accordance with Good Publication Practice (GPP) guidelines (www.ismpp.org/gpp-2022), and was funded by GSK and Vir Biotechnology.

## Authorship

All named authors meet the International Committee of Medical Journal Editors (ICMJE) criteria for authorship for this article, take responsibility for the integrity of the work as a whole and have given their approval for this version to be published.

## Author contributions

MD, VP, HT, HB, EL and AS were involved in the study design, conception and execution. HT, DCG and CM took part in the acquisition and analysis of data. All authors were involved in data interpretation; drafting, revising or critically reviewing the manuscript; gave final approval of the version to be published; have agreed on the journal to which the article has been submitted; and agree to be accountable for all aspects of the work.

## Compliance with ethics guidelines

This study received data access and processing approval from the Public Benefit and Privacy Panel for Health and Social Care, NHS Scotland on 9 November 2022. We complied with all applicable laws regarding subject privacy. No direct subject contact or primary collection of individual human subject data occurred as part of this study. Study results were reported in tabular form and aggregate analyses that omitted subject identification. Publications, reports or any other research outputs do not include subject identifiers and low patient numbers were suppressed.

